# Exploring Machine Learning Strategies in COVID-19 Prognostic Modelling: A Systematic Analysis of Diagnosis, Classification and Outcome Prediction

**DOI:** 10.1101/2024.03.17.24304450

**Authors:** Reabal Najjar, Md Zakir Hossain, Khandaker Asif Ahmed, Md Rakibul Hasan

## Abstract

**Background:** The COVID-19 pandemic, which has impacted over 222 countries resulting in incalculable losses, has necessitated innovative solutions via machine learning (ML) to tackle the problem of overburdened healthcare systems. This study consolidates research employing ML models for COVID-19 prognosis, evaluates prevalent models and performance, and provides an overview of suitable models and features while offering recommendations for experimental protocols, reproducibility and integration of ML algorithms in clinical settings.

**Methods:** We conducted a review following the PRISMA framework, examining ML utilisation for COVID-19 prediction. Five databases were searched for relevant studies up to 24 January 2023, resulting in 1,824 unique articles. Rigorous selection criteria led to 204 included studies. Top-performing features and models were extracted, with the area under the receiver operating characteristic curve (AUC) evaluation metric used for performance assessment.

**Results:** This systematic review investigated 204 studies on ML models for COVID-19 prognosis across automated diagnosis (18.1%), severity classification (31.9%), and outcome prediction (50%). We identified thirty-four unique features in five categories and twenty-one distinct ML models in six categories. The most prevalent features were chest CT, chest radiographs, and advanced age, while the most frequently employed models were CNN, XGB, and RF. Top-performing models included neural networks (ANN, MLP, DNN), distance-based methods (kNN), ensemble methods (XGB), and regression models (PLS-DA), all exhibiting high AUC values.

**Conclusion:** Machine learning models have shown considerable promise in improving COVID-19 diagnostic accuracy, risk stratification, and outcome prediction. Advancements in ML techniques and their integration with complementary technologies will be essential for expediting decision-making and informing clinical decisions, with long-lasting implications for healthcare systems globally.

## 1 Introduction

Since the first cluster of cases was identified in Wuhan, China, in December 2019, the novel coronavirus—SARS-CoV-2, the causative agent behind the coronavirus disease 2019 (COVID-19)—has taken the world by storm. Declared a global pandemic by the World Health Organisation (WHO) on March 11, 2020, COVID-19 has led to significant health and economic losses [1]. As of 11 March 2023, exactly three years since officially recognised as a pandemic, SARS-CoV-2 has caused more than 676,609,955 cases and claimed 6,881,955 lives in 222 countries [2].

The COVID-19 pandemic has profoundly affected the global landscape, leaving an enduring mark on healthcare systems, economies, and societies worldwide. Since the onset of the outbreak, healthcare professionals have diligently worked to contain the virus and mitigate its effects. Despite the implementation of public health measures aimed at controlling the spread of COVID-19, many countries continue to grapple with recurrent outbreaks, causing significant disruptions to healthcare services and society at large. The recent Omicron variant outbreak has placed an additional burden on testing, medical center capacity, and human resources. It is not unlikely that such outbreaks will be witnessed in future waves of COVID-19 worldwide. Although many national governments have lifted COVID-19 restrictions, scientists warn that the endemic circulation of SARS-CoV-2, possibly accompanied by seasonal epidemic peaks, is likely to continue to cause significant disease burden [3].

Reverse transcription-polymerase chain reaction (RT-PCR) is still regarded as the gold standard for diagnosing COVID-19 infections. However, it has several significant drawbacks, such as extended turnaround times, scarcity of reagents, false-negative rates up to 20%, and the requirement for accredited laboratories, expensive equipment, and trained specialists [4]. Consequently, RT-PCR may be more appropriately considered a primary detection measure rather than a diagnostic standard. Rapid antigen tests (RATs) have gained popularity for large-scale testing due to their cost-effectiveness and ability to deliver results rapidly; however, their sensitivity relies on the skill and expertise of the individual administering the test [5]. As a result, the need for innovative solutions has become more pressing than ever.

Recently, novel solutions such as artificial intelligence have experienced a significant upsurge, which holds the potential to strengthen worldwide initiatives in addressing the COVID-19 crisis. Artificial intelligence, commonly abbreviated as AI, is a field of computer science that applies computational algorithms to replicate human intelligence. In other words, the term ‘AI’ refers to a device’s ability to mimic our own cognitive function. This imitation is achieved through pattern recognition and has the potential to minimise the need for human intervention in repetitive and mundane tasks. AI is becoming increasingly pervasive in various industries, from autonomous electric cars to virtual assistants like Alexa, and from predictive analysis of the stock market to medical diagnosis. Its uses have become imperative in different fields, including the clinical sector [6]. As AI continues to evolve, it has the potential to revolutionise several industries, from manufacturing to healthcare, by increasing efficiency and accuracy while reducing human error.

In conjunction with AI, machine learning (ML) is frequently employed in clinical environments as a subset of AI that focuses on the autonomous enhancement of computer algorithms through iterative learning processes, including training, validation, and testing on various datasets. Presently, ML models are at the forefront of the AI domain. These models excel at identifying patterns within extensive datasets, enabling the development of sophisticated reasoning systems for patient risk categorisation and more informed decision-making processes. ML methods offer the advantage of harnessing richer clinical data for predictive modeling, an approach that has been widely investigated throughout the COVID-19 pandemic.

Employing data mining techniques and adaptive ML algorithms has consistently demonstrated superior performance in comparison to traditional statistical methods, accurately forecasting individual patient outcomes and effectively modeling disease prognosis and risk assessment. Additionally, research has shown that ML algorithms can outperform medical professionals in certain radiological diagnostic tasks [7, 8]. These findings highlight the need for developing models that integrate multiple data sources, such as radiological imaging, clinical manifestations, and laboratory test results, to enhance diagnostic accuracy. By leveraging ML algorithms and diverse data modalities, more accurate and reliable diagnostic models can be established, ultimately leading to improved patient outcomes and more efficient pandemic control. These tools can be employed by decision-makers and public health authorities to design and implement policies and actionable measures that are particularly critical during challenging times, such as those posed by the global COVID-19 pandemic.

Hence, developing tailored ML models for COVID-19 can aid in assessing the disease’s prognosis. These approaches offer several benefits, including high diagnostic accuracy, effective severity stratification, and reliable outcome prediction due to their remarkable sensitivity and specificity, exceptional consistency and repeatability, and the potential for large-scale deployment. This makes ML an essential tool in the ongoing battle against COVID-19 and the optimisation of healthcare resource allocation.

### 1.1 Related Work

In the last three years, there has been a substantial increase in publications related to the application of ML for COVID-19. A multitude of studies has employed prognostic variables to leverage ML to address and overcome the myriad of challenges confronted by healthcare professionals during the pandemic.

In their scoping review, Abd-Alrazaq and colleagues examine the role of AI in combating the COVID-19 pandemic. They identified 219 relevant articles and found that AI has played a significant role in diagnosing COVID-19, forecasting the virus’s spread, developing risk models for patient outcomes, contact tracing, public health management, and drug development. The authors highlight the need for interdisciplinary collaborations, data sharing, and addressing ethical, legal, and social implications [9].

The scoping review by Naseem and colleagues investigates the possibilities of utilising AI and ML to address the COVID-19 pandemic. The authors identified various algorithm applications in areas such as diagnosis, treatment, and prevention, as well as their potential to enhance healthcare systems. The study highlights the significant role that ML can play in combating COVID-19 and underscores the need for further research to maximise their benefits in resource-limited settings [10].

Chee et al. reviewed nineteen studies on AI applications in COVID-19 management within intensive care and emergency settings. The findings reveal the potential of AI to enhance patient care and resource allocation, with some studies reporting diagnostic accuracy as high as 96% and AUC (area under the receiver operating characteristic curve) values up to 0.98 for prognosis [11].

The systematic review by Wang et al. investigated eighty-seven studies on AI applications in addressing COVID-19, with a focus on diagnosis, treatment, prevention, and public health management. The findings reveal AI’s potential, particularly in medical imaging-based diagnosis, where some studies reported sensitivity and specificity values exceeding 90%. Despite these promising results, the study emphasises the need for additional research, validation, and standardisation to ensure AI’s successful integration into clinical and public health settings [12].

Mulrenan et al.’s article review the use of ML models for COVID-19 diagnosis through CT scans and chest X-rays. While the authors report promising results in accurately identifying COVID-19 cases, they also raise concerns about small dataset samples and poor external validity, stemming from the limited inclusion of features or variables in the examined studies. The findings emphasise the need for additional research, validation, and standardisation to ensure reliable AI-based diagnostic tools for improved detection and management of the disease [13].

Numerous prognostic scores and models suffer from inherent biases due to inadequate reporting and methodology. The most common limitations of these studies include the reliance on individual features for risk assessment and outcome prediction, as well as insufficient sample sizes and limited datasets, which can be attributed to the difficulties of collecting clinical data amid a pandemic. Such an approach, however, introduces bias, diminishes external validity, and limits the transferability to clinical practice, ultimately compromising precision and power.

### 1.2 Aims and Objectives

To address a gap in the scientific literature, we conducted a comprehensive systematic literature review to investigate the prognostic potential of ML models in facilitating automated detection, severity classification, and outcome prediction set against the backdrop of COVID-19. The goal of this analysis is not to determine causality, but rather to select a model that performs best for the given dataset and prediction task. Our approach entailed a critical appraisal of a broad range of demographic, clinical, laboratory, and imaging features utilised in conjunction with ML algorithms. As such, this study aimed to:

1. *Present a thorough synthesis of pertinent studies that employed ML models for forecasting COVID-19 prognosis, providing an exhaustive overview of data sources, ML methodologies, types of features, and associated results*.
2. *Evaluate and compare the most frequently utilised ML models in COVID-19 prognostication by examining performance metrics and identify the most prevalent features and datasets used in these models*.
3. *Offer an overview of the ML models best suited for each specific feature category, along with recommendations for reporting experimental protocols and results to facilitate reproducibility*.

As a result, the application of ML to various modalities of clinical and non-clinical data has often yielded high diagnostic accuracies in patients, thereby fostering more accurate and informed decision-making by encouraging the integration and adoption of ML algorithms in clinical settings.

### 1.3 Study Structure

In the following section, we elaborate on the methodology employed in our literature review, outlining the search strategy utilised to identify the most relevant studies. We chose studies adhering to the inclusion criteria to ascertain the most efficient ML models and most accurate clinical datasets to prognosticate the disease and its progression. Additionally, we discuss the data extraction process and the performance metrics utilised for evaluation.

The subsequent results section presents the outcomes of our search criteria, highlighting the final number of included studies. We provide a summary of the studies’ key characteristics, objectives, the primary features utilised, and their main categories, as well as the most frequently utilised ML models.

In the final primary section, we contextualise our findings within three central prognostic themes: (I) automated diagnosis, (II) severity classification, and (III) outcome prediction. We explore successful clinical scenarios where the application of robust and validated ML methods proves effective for each theme. A synthesis and outlook encompassing the impact of ML models in terms of robustness and transferability is provided in the discussion. Furthermore, we recognise recent accomplishments, advancements, and emerging challenges encountered in adopting this state-of-the-art technology while also discussing potential avenues for future research investigations.

## 2 Methods

Our systematic literature review adhered to the guidelines outlined in the PRISMA (Preferred Reporting Items for Systematic Reviews and Meta-Analyses) framework for its preparation and reporting [14]. The primary aim of this review is to critically examine the current literature on ML utilisation for COVID-19 to find the most efficient models for disease prediction and identify the most representative predictors for diagnosis, severity classification, and outcome prognostication.

All data generated or analysed during this study are included in this published article and its supplementary information files.

### 2.1 Search Strategy

Five academic databases were searched up to 24 January 2023: Google Scholar, Web of Science, PubMed, Scopus, and MEDLINE. To find relevant studies on the use prognostic use of AI/ML models in COVID, the following search string was used: “COVID*”, “SARS-CoV-2”, “artificial intelligence”, “AI”, “machine learning”, “ML”, and “prognos*” (where * denotes wildcard characters).

### 2.2 Selection Criteria

After applying the search criteria to the selected libraries, a total of 2,721 studies from five scientific databases were retrieved. 1,824 unique studies remained after duplicates were removed. To be included in our research, studies had to fulfil the following rigorous selection criteria:

Inclusion criteria include: (1) *Studies that explicitly addressed the COVID-19 issue;* (2) *applied at least one AI/ML algorithm;* (3) *were original, peer-reviewed research articles; and* (4) *were written and published in English only*.

Exclusion criteria include: (1) *Studies published before 2019;* (2) *discussed AI/ML algorithms but did address COVID-19;* (3) *mentioned COVID-19 but did not utilise AI/ML algorithms; and* (4) *reviews, surveys, case reports, extended abstracts, or conference posters*.

These criteria ensured that the selected studies were relevant to our research and met a certain standard of quality, providing a robust foundation for our analysis and conclusions. To guarantee that the studies met the criteria, they underwent a thorough review process. First, the titles were screened, followed by a subsequent screening of the abstracts. Finally, an independent reviewer conducted a full-text review of the studies that passed the initial screening. Any discrepancies in the reviewer’s assessments were resolved through discussion and, if necessary, by an additional reviewer.

### 2.3 Data Extraction

To address our research questions, we collected relevant qualitative and quantitative data from selected studies using a data extraction form. For each of the 204 included studies, we extracted and recorded the following data: basic information (title, authors, year of publication), study aim and major findings, prognostic theme, patient population, feature category, performance metrics, highest performing model, and most representative feature. We analyzed the extracted data to gain insights into the current state of research on machine learning (ML) and COVID-19 by presenting the findings in the form of tables and figures.

### 2.4 Performance Metrics

For each study, we identified the ML model that achieved the highest performance metric and the associated feature. Suppose a study used only one model, that was considered the selected model. There was significant variation among the studies in terms of the characteristics, outcome measures, and model performance. Therefore, we did not attempt to calculate a point estimate for the overall models’ performance. However, since there are numerous models, we analyzed their predictive features and performance by examining the AUC (area under the receiver operating characteristic curve), which is a summary measure of sensitivity and specificity. While no single performance metric gives an overall representation, the AUC was decisively selected as the main performance metric as 181 out of 204 included studies (88.7%) reported the AUC. Among the remaining 23 studies that did not report AUC, accuracy was used to evaluate performance.

## 3 Results

### 3.1 Literature Review

Our search identified 2,721 studies, of which 1,824 were unique. After initial screening based on title, 626 studies remained, with a further 354 studies excluded after the abstract screening. The remaining 272 studies underwent a full-text review, during which 68 studies were excluded after a thorough examination. Ultimately, a total of 204 full-text studies investigating the prognostic use of machine learning (ML) models for COVID-19 were incorporated into our review. The reporting process adhered to the PRISMA flow diagram guidelines (Figure 1).

**Figure 1:**
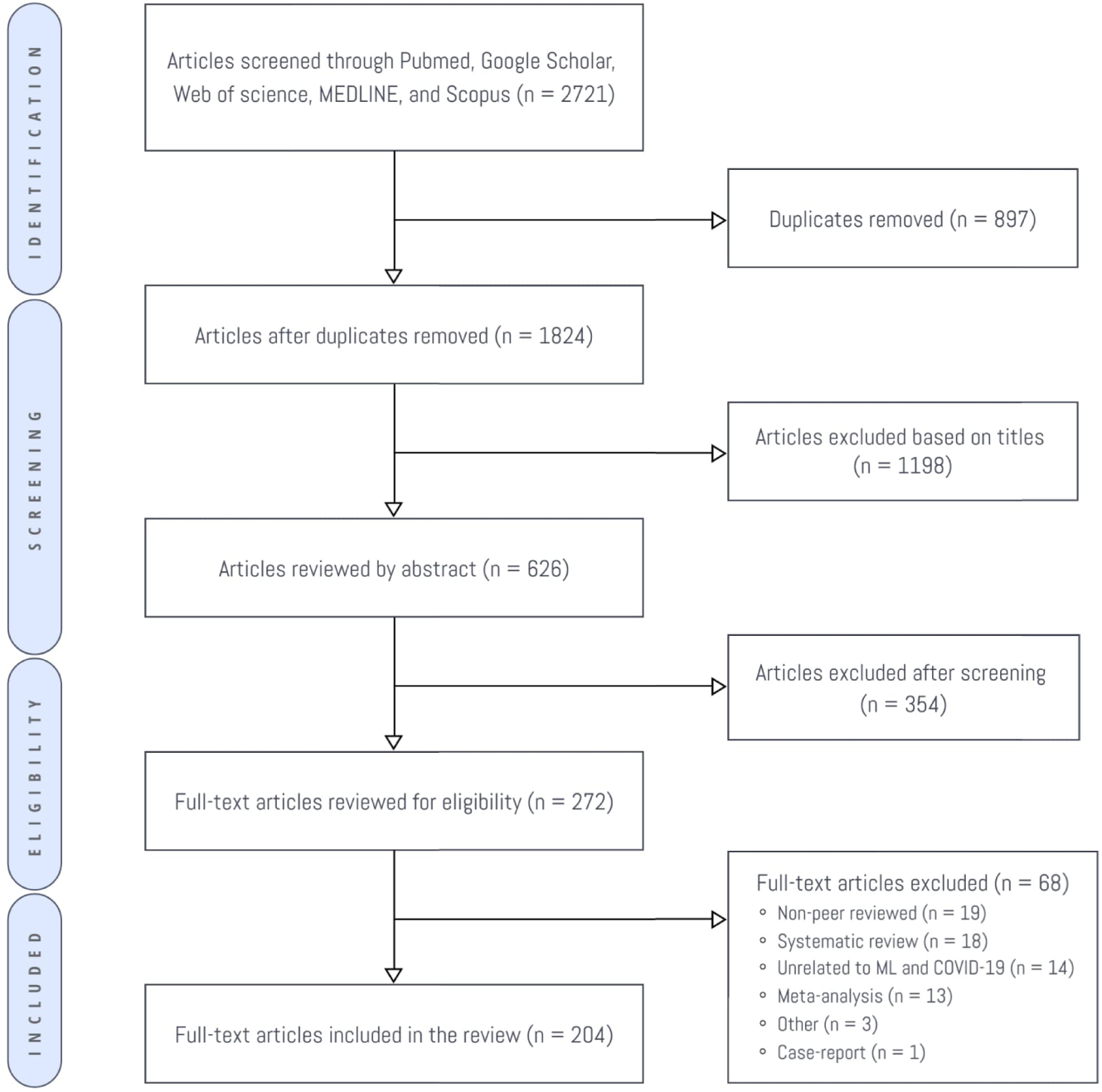
Flow chart depicting the study selection process in accordance with the PRISMA guidelines

### 3.2 General Characteristics

The studies included in this analysis represent a diverse and geographically extensive dataset, derived from 29 distinct countries across various continents (Figure 2). The United States stood out as the most represented, contributing 50 studies (24.5%), followed by China with 45 studies (22.1%), Italy with 16 studies (7.8%), Iran with 13 studies (6.4%), and India with 10 studies (4.9%). The countries reflect areas most affected by the pandemic in early 2020. Additionally, this global representation highlights the widespread interest and collaborative efforts in leveraging ML techniques to combat COVID-19.

**Figure 2:**
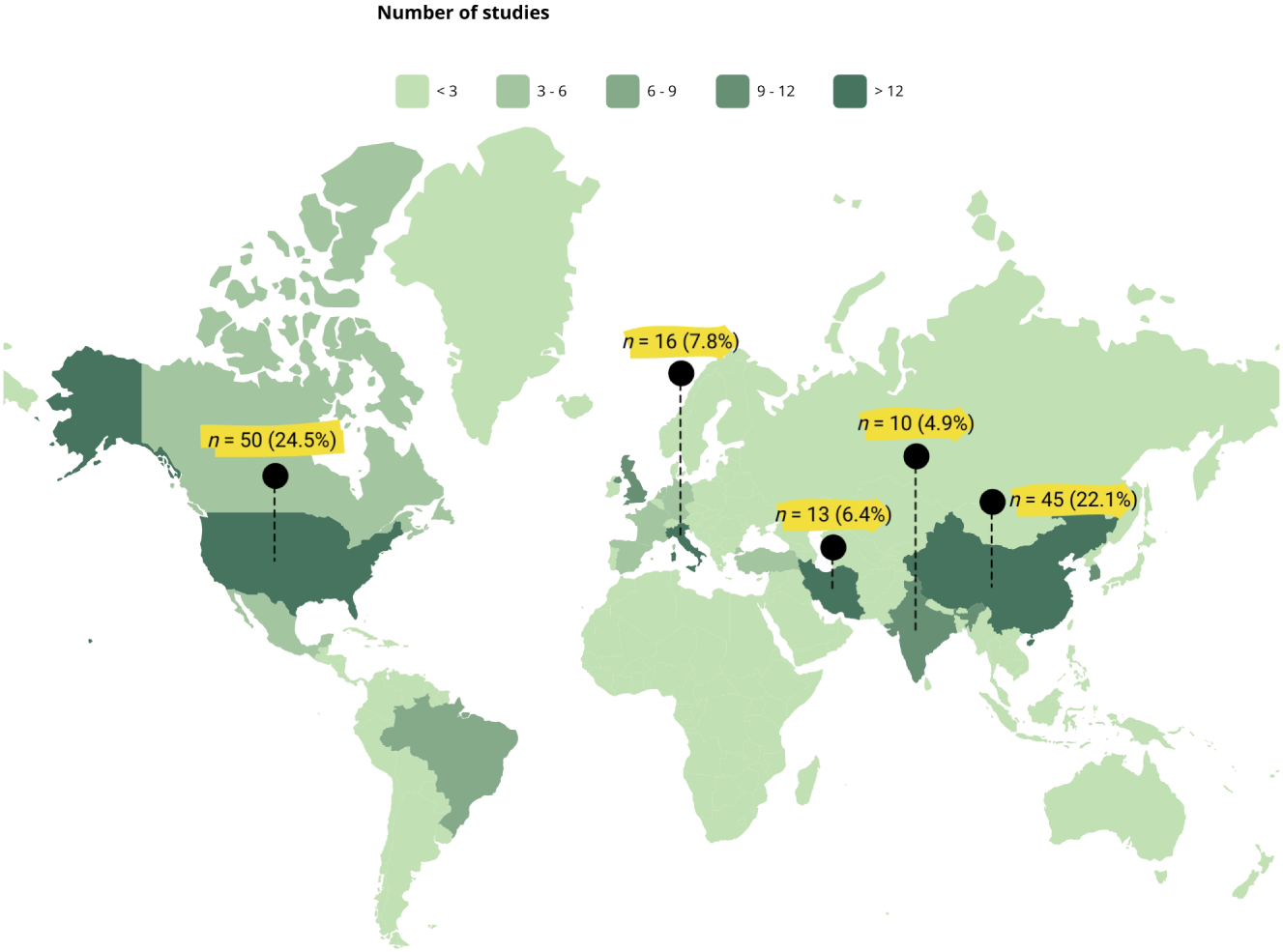
Geographical distribution of published studies by country, with top five highlighted

The included studies were all published from 2020 onward, demonstrating a significant increase in the annual number of papers in this field. This surge in research activity highlights the pressing need to find effective solutions to the challenges posed by the pandemic. ML has risen to prominence as a formidable instrument in combating COVID-19, enabling the analysis of vast amounts of data and the identification of complex patterns that would be impossible for humans to detect. Its ability to rapidly process and interpret data has been crucial in informing decision-making processes, predicting outbreaks, and developing effective treatments and vaccines. As such, it has become an indispensable tool in the ongoing battle against the pandemic.

The 204 studies analyzed exhibited a diverse range of patient sample sizes, signifying the broad scope of research conducted within this domain. The average sample size was 4,943.6, with a median of 625. The smallest patient sample size consisted of only 44 participants, while the largest sample size included an impressive 263,007 patients. This disparity illustrates the wide spectrum of study sizes and the rapidly growing interest in this area of research. Moreover, the variation in sample sizes underscores the adaptability of ML models to different datasets, thereby emphasising their potential for widespread clinical application.

### 3.3 Performance Metrics

The evaluation of ML models is a critical aspect of the development process, and a range of metrics have been devised to assess their performance. In the present study, all 204 articles analyzed specified an outcome measure or metric to evaluate the performance of their ML models.

The most commonly used metric among these studies was AUC, which was used to evaluate the performance of each model in 178 out of 204 studies (87.3%). AUC was used either independently in 156 studies or in combination with accuracy in 22 studies. Accuracy was used exclusively in 23 studies (11.3%). The remaining 3 studies used performance metrics other than AUC and/or accuracy, such as precision (*n* = 1), coefficient of determination or R2 (*n* = 1), and recall (*n* = 1).

The widespread use of AUC in ML studies can be attributed to its ability to measure the model’s ability to discriminate between positive and negative samples across different thresholds. Its use, in combination with accuracy, can provide a more comprehensive assessment of the model’s performance. This is particularly important in applications such as medical diagnosis, where the cost of false positives and false negatives can be significant. Furthermore, the AUC provides a comprehensive assessment of the model’s performance by taking into account all possible threshold values, providing a single value that summarises its discriminatory power.

### 3.4 Feature Groups

In order to enhance the clarity of our discussion, we will refer to the predictive variables or datasets utilised for constructing the studies’ algorithms as “features.” These features were systematically classified into five distinct categories, each providing valuable information for COVID-19 diagnosis, classification, and progression prediction.

#### 3.4.1 Demographic Characteristics

Demographic features such as age, body mass index (BMI), gender, and length of hospital stay offer valuable information about patients’ vulnerability to COVID-19 and possible disease outcomes. Age above 60 years has been consistently reported as a significant risk factor for severe infection and mortality (*n* = 34, 16.7%). Gender, albeit less frequently investigated (*n* = 1, 0.5%), has also been implicated in influencing disease severity, with males often experiencing worse outcomes. BMI (*n* = 2, 0.98%) plays a role in identifying individuals with obesity, who are at a higher risk for complications. Length of hospital stay (*n* = 1, 0.5%) can provide insights into the overall burden on healthcare systems and prognostic implications for patients.

#### 3.4.2 Co-morbid Conditions

The manifestation of pre-existing medical conditions, such as chronic obstructive pulmonary disease (COPD) and asthma, results in unfavourable consequences due to compromised pulmonary function and heightened susceptibility to respiratory infections [15, 16]. Our review included only one study for each condition (*n* = 1, 0.5%), highlighting the need for further investigation to gain a comprehensive understanding of the influence of these co-existing medical conditions on the prognosis of COVID-19. Other co-morbid conditions, such as diabetes and cardiovascular diseases, have also been linked to worse outcomes and warrant further investigation.

#### 3.4.3 Vital Signs

Oxygen saturation (SpO2) is a vital physiological parameter that indicates the percentage of oxygenated haemoglobin in the blood. This metric serves as a crucial indicator for monitoring a patient’s respiratory function and overall health status, underscoring its importance as a tool for patient care and clinical decision-making. Lower SpO2 levels indicate potential hypoxaemia, a condition prevalent in severe COVID-19 cases. Our review identified 18 studies (8.8%) that emphasised the importance of SpO2 in predicting COVID-19 severity, progression, and patient outcomes.

#### 3.4.4 Clinical Symptoms

Altered mental status (AMS), cough, and tachypnoea were the most common clinical features employed in the studies (*n* = 2 each, 0.98%). AMS can indicate neurological involvement in COVID-19, while cough and tachypnoea reflect respiratory distress. Dyspnoea, fever, cough, and sore throat, although less frequently investigated, are also classic symptoms of COVID-19 and may hold predictive value for disease severity and progression.

#### 3.4.5 Chest Imaging

Both chest radiography (CXR) and chest computed tomography (CT) scans provide valuable insights into lung involvement, which is crucial in the assessment of COVID-19 patients. CXR (*n* = 26, 12.7%) offers a rapid, low-cost, and accessible method for detecting lung abnormalities, while CT scans (*n* = 45, 22.1%) provide higher resolution images, enabling more accurate identification of lung lesions and disease extent.

#### 3.4.6 Serum Biomarkers

Blood-based biomarkers offer a non-invasive approach to assess the severity and progression of COVID-19. Inflammatory markers such as lactate dehydrogenase (LDH), the neutrophil-to-lymphocyte ratio (NLR), and C-reactive protein (CRP) indicate the presence of inflammation and tissue damage. Immune parameters, including cytokines like interferon and interleukin-6 (IL-6), provide insights into the immune response to infection. Blood coagulation parameters (ferritin, D-dimer, and prothrombin) may reveal potential coagulation abnormalities, while biochemical markers such as the glomerular filtration rate (GFR), creatinine, and aspartate aminotransferase (AST) shed light on kidney and liver function. The most frequently used features in the studies were LDH and NLR (*n* = 16 each, 7.8% each), CRP (*n* = 11, 5.4%), and cytokines and D-dimer (*n* = 3 each, 1.5%). By incorporating these diverse serum biomarkers into ML models, researchers can gain a comprehensive understanding of the complex interplay between host response, systemic inflammation, organ function, and coagulation abnormalities in COVID-19 patients.

Through a meticulous analysis of various feature categories, our systematic review endeavors to advance the current knowledge of the possible applications of ML in predicting the progression, diagnosis, and classification of COVID-19. By thoroughly exploring these different categories of features, we aim to provide valuable insights into the potential of ML in this domain.

### 3.5 Machine Learning Models

This systematic review conducted an in-depth examination of methodological approaches and performance outcomes for each ML model presented in the studies. In instances where multiple models were introduced within a single study, only the top-performing technique was surveyed. A total of 204 ML algorithms were examined, from which 21 unique models were identified. Convolutional Neural Networks (CNN) emerged as the most prevalent model, appearing in 51 studies, followed by eXtreme Gradient Boosting (XGB) in 39 studies, and Random Forest (RF) in 34 studies. The 21 models were classified into six categories (Figure 3).

**Figure 3:**
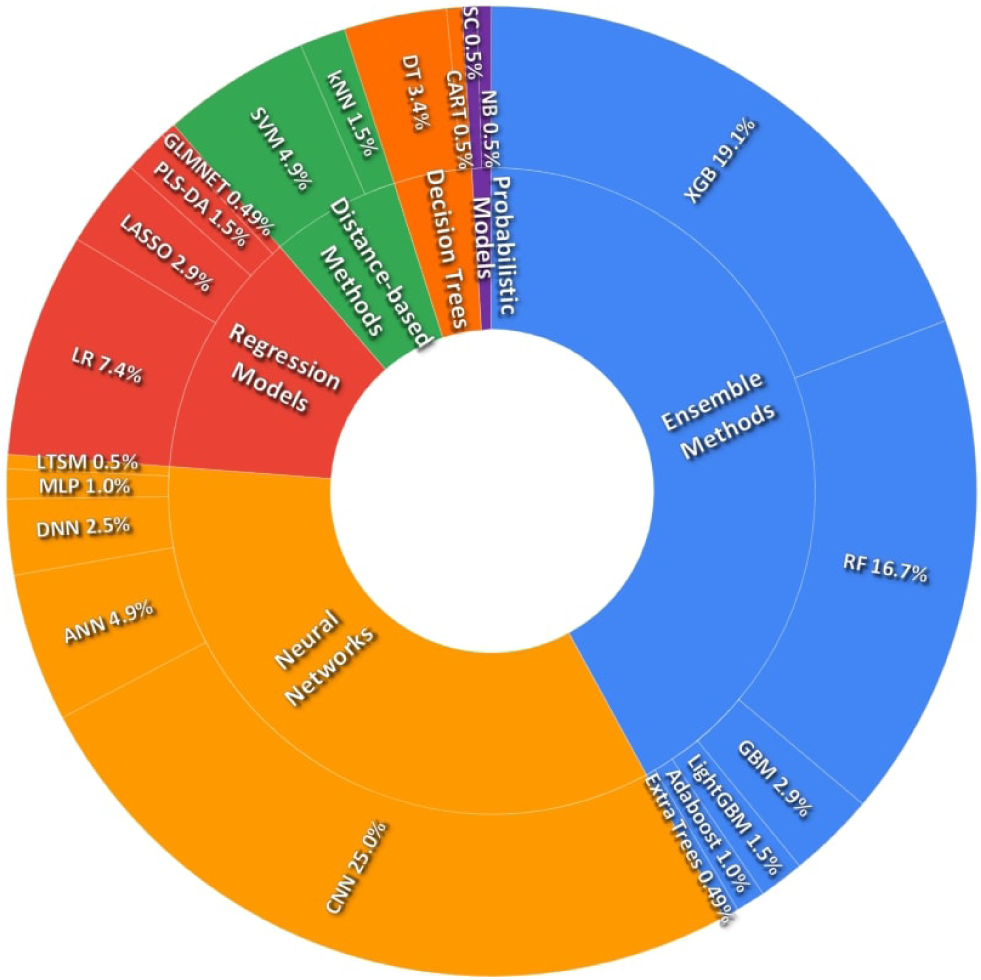
Sunburst chart: Machine learning categories and models in COVID-19 research

#### 3.5.1 Neural Networks

Neural networks (*n* = 76, 37.3%) comprise a collection of algorithms that emulate the structure and function of biological neural networks. These models demonstrate remarkable proficiency in discerning intricate patterns and representations from vast datasets, making them ideal for performing tasks that require high-level image and speech recognition, natural language processing, as well as biomedical applications [17]. This category includes CNN (*n* = 51, 25%), Artificial Neural Networks (ANN; n = 10, 4.9%), Deep Neural Networks (DNN; *n* = 5, 2.5%), Multilayer Perceptron (MLP; *n* = 2, 0.98%), and Long Short-Term Memory networks (LSTM; *n* = 1, 0.49%).

#### 3.5.2 Ensemble Methods

Ensemble methods (*n* = 85, 41.7%) integrate multiple base models to enhance generalisation and predictive performance. This is achieved by leveraging the strengths of individual models, which often have different strengths and weaknesses, to generate a more accurate and robust aggregated model [18]. This category comprises XGB (*n* = 39, 19.1%), RF (*n* = 34, 16.7%), Gradient Boosting Machines (GBM; *n* = 6, 2.9%), LightGBM (*n* = 3, 1.5%), Adaptive Boosting (Adaboost, *n* = 2, 0.98%), and Extra Trees Classifier (*n* = 1, 0.49%).

#### 3.5.3 Decision Trees

Decision trees (*n* = 8, 3.9%) utilise tree-like structures that recursively partition input data into subsets based on feature values. This allows for the management of both categorical and continuous data, making them highly versatile. Furthermore, decision trees offer a high level of interpretability, making them an attractive option for those seeking easy implementation and comprehension of their results [19]. This category consists of Decision Trees (DT, *n* = 7, 3.4%) and Classification and Regression Trees (CART; *n* = 1, 0.49%).

#### 3.5.4 Regression Models

Regression models (n = 25, 12.3%) are statistical approaches that enable researchers to examine and model the association between a dependent variable and one or more independent variables. These models can take the form of either linear or nonlinear regressions, and they are commonly leveraged to make predictions or projections in a variety of contexts [20]. This category comprises Logistic Regression (LR; *n* = 15, 7.4%), Least Absolute Shrinkage and Selection Operator (LASSO; *n* = 6, 2.9%), Partial Least Squares Discriminant Analysis (PLS-DA; *n* = 3, 1.5%), and Generalised Linear Model with Elastic Net Regularisation (GLMNET; *n* = 1, 0.49%).

#### 3.5.5 Distance-based Methods

Distance-based methods (*n* = 15, 7.4%) are predicated on the concept of distance or similarity between data points to make predictions. These techniques are particularly adept at handling tasks that encompass high-dimensional data or complex associations [21, 22]. This category includes Support Vector Machines (SVM; *n* = 12, 4.9%) and k-Nearest Neighbors (kNN; *n* = 3, 1.5%).

#### 3.5.6 Stochastic Models

Stochastic models (*n* = 2, 0.98%) compute the probability distribution in a feature dataset, providing a quantification of the inherent uncertainty associated with the ensuing predictions [23]. Often employed in applications where probabilistic interpretations are vital, such as medical diagnosis and risk assessment, this category encompasses Naïve Bayes (NB; *n* = 1, 0.49%) and Shrunken Centroids (*n* = 1, 0.49%).

### 3.6 Prognostic Themes

To delineate the various goals and objectives of the studies included in this analysis, we categorised them into groups according to their ML prognostication model and overall purpose. We identified three major prognostic themes: (I) *automated detection and AI-assisted diagnosis*, (II) *severity classification and risk stratification*, and (III) *prediction of outcome and disease progression* (4).

#### 3.6.1 Automated detection and ML-assisted diagnosis

Automated detection and diagnosis of diseases are essential for timely and accurate medical interventions, particularly in the context of a pandemic. The implementation of AI into traditional diagnostic approaches could potentially lead to swifter, more definitive, and more precise determinations for self-isolation guidelines and the identification of high-risk patients. In our systematic review, we analysed 37 studies (18.1%) that primarily focused on detecting and diagnosing COVID-19 in patients with suspected infection using ML techniques. These studies showcased the integration of imaging data, such as CXR and CT scans, along with clinical and laboratory data to develop sophisticated diagnostic models (see Supplementary Table 1 for an overview of studies utilising ML for COVID-19 automated detection).

In the context of automated detection of COVID-19, the most frequently used ML models were CNN, GBM, and RF/ANN. CNN was the most widely used model, with 20 out of 38 studies (52.7%) employing it for diagnostic applications. GBM and RF/ANN were used in 10.5% and 7.9% of studies, respectively. Furthermore, our review also revealed the top three features used in this category. Chest CT was the most commonly used feature, with 34.2% of studies, followed by CXR as the second most commonly used feature, with 23.7% of studies utilising it. Finally, NLR was used by 10.5% of studies (Figure 5a).

**Figure 4:**
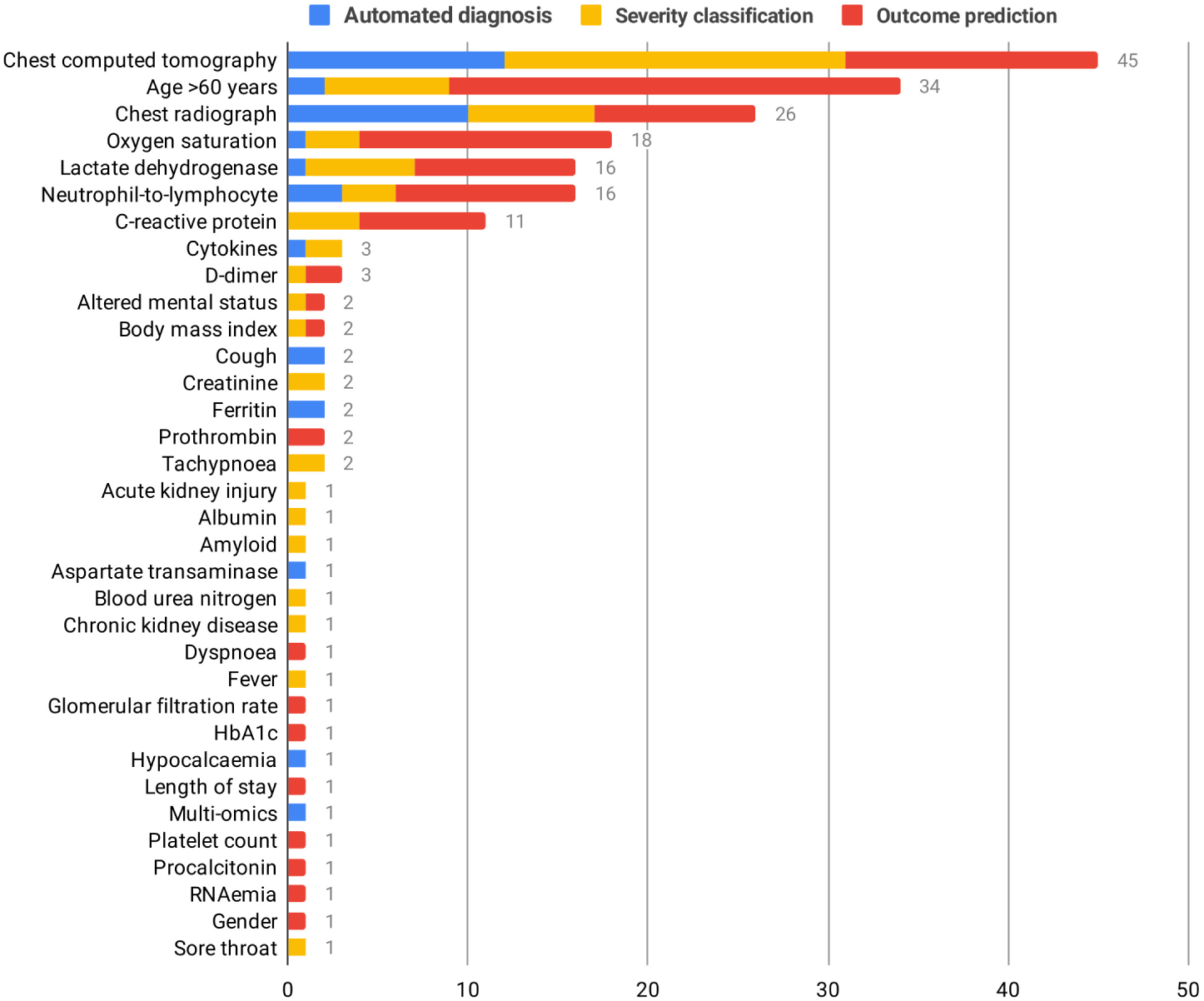
Frequency distribution of top-performing features and prognostic themes

**Figure 5:**
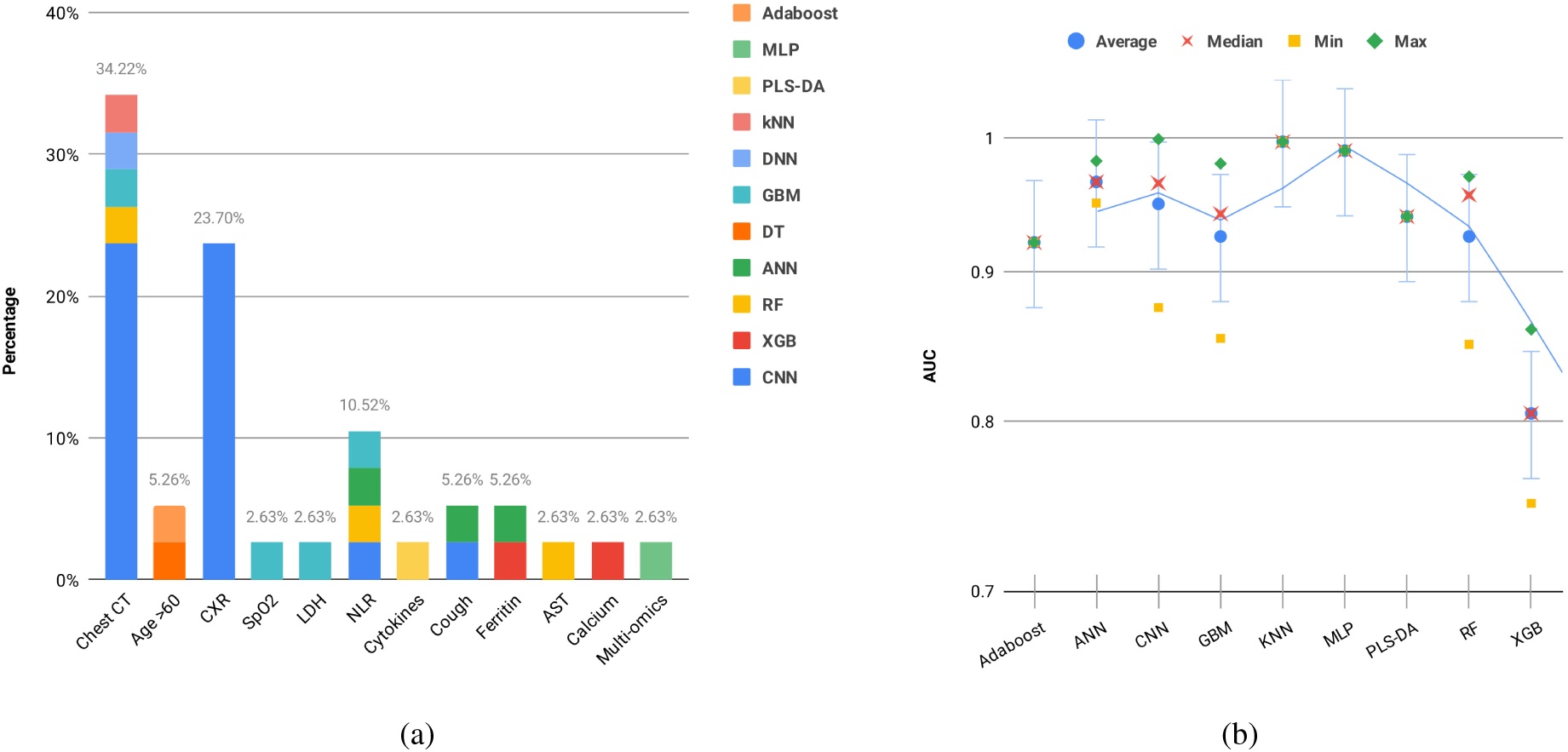
(a) Percentage of machine learning models applied per feature in automated diagnosis and (b) Performance metrics for machine learning models in automated diagnosis

In our analysis of various ML models, the top three performers were kNN, MLP, and ANN, with average AUC values of 0.997, 0.990, and 0.966, respectively (Figure 5b).

#### 3.6.2 Severity Classification and Risk Stratification

Artificial intelligence has applications beyond merely formulating a diagnosis; it can also make predictions about disease quantification and staging for individuals who are already infected. A clinical algorithm that accurately predicts and stratifies patients with a higher likelihood of experiencing adverse outcomes is crucial for optimising healthcare resource allocation for those requiring urgent care. By identifying individuals at greater risk for severe illness, complications, or mortality, healthcare providers can prioritise early intervention and targeted treatment strategies for high-risk patients. Disease prediction has been extensively studied using well-established ML algorithms for handling binary or multi-class learning problems. Classifying patients into predefined groups allows for the development of predictive models that assess risk stratification with generalisable performance (see Supplementary Table 2 for an overview of studies utilising ML for COVID-19 severity classification).

We employed the WHO Clinical Progression Scale, a modified assessment tool devised to gauge the severity of patient illness during infectious disease outbreaks by leveraging readily available clinical information [24]. In this regard, various studies on COVID-19 severity classification using traditional algorithms and deep learning methods were included in our analysis (*n* = 65, 31.7%). Among these models, CNN, XGB, and RF/SVM stand out as the most frequently used, with 26.2%, 18.5%, and 12.3% usage, respectively. The top three features utilised in these studies include chest CT (29.2%), age *≥* 60 years/CXR (10.8%), and LDH (9.2%), which are demonstrated in Figure 6a. In our performance evaluation, the leading ML models were as follows: kNN with an average AUC of 0.980, ANN with an average AUC of 0.935, and XGB with an average AUC of 0.916 (Figure 6b).

**Figure 6:**
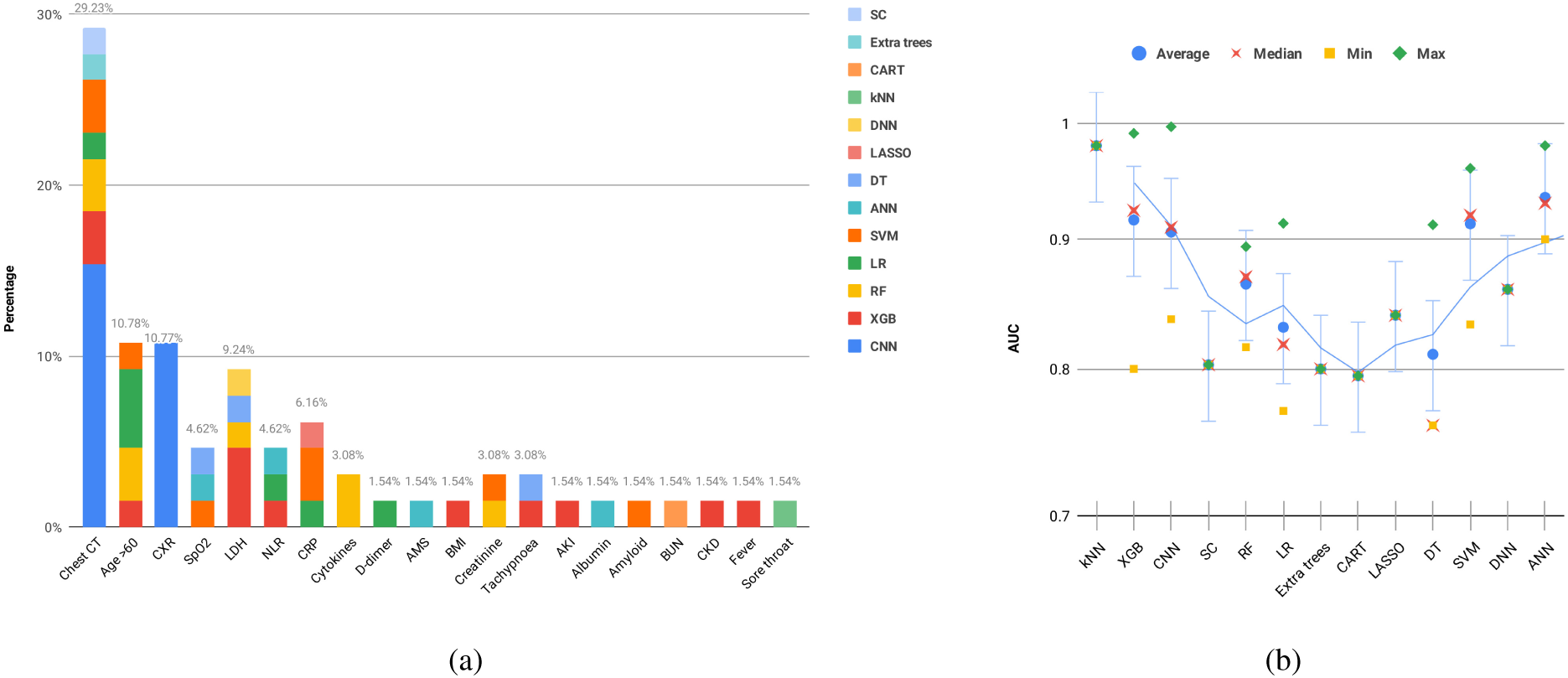
(a) Percentage of machine learning models applied per feature in severity classification and (b) Performance metrics for machine learning models in severity classification

#### 3.6.3 Prediction of Outcomes and Disease Progression

The initial manifestations of COVID-19 are typically mild, with a considerable percentage of patients exhibiting no overt symptoms prior to the onset of respiratory failure. From a clinical perspective, this poses a challenge in forecasting the progression of disease severity in patients until the development of respiratory failure becomes apparent. Early outcome prediction through non-invasive methods can provide valuable information on the optimal timing, location, and target population for effective interventions. Such models have the potential to establish robust frameworks for diverse medical tasks and enhance patient triage processes.

By accurately predicting patient outcomes through AI algorithms, healthcare providers can initiate early intervention and optimise the use of healthcare resources. In this review, we focused on studies that sought to forecast patient prognosis and disease progression with respect to outcomes, such as ICU admission, mechanical ventilation, and mortality risk. Precisely half of the studies incorporated in our review (*n* = 102) aimed to predict patient outcomes by employing ML models (see Supplementary Table 3 for an overview of studies utilising ML for COVID-19 outcome prediction).

The top three ML models employed in these studies were XGB (*n* = 25, 24.5%), RF (*n* = 23, 22.5%), and CNN (*n* = 14, 13.7%). Each of these models has its unique strengths and capabilities, contributing to their popularity in the field. Moreover, we identified the top three features critical for predicting patient outcomes. Age *≥* 60 (*n* = 25, 24.5%) was the most common feature, signifying that older patients have a higher risk of severe outcomes. The second most frequent feature was SpO2/Chest CT (*n* = 14 each, 13.7%), highlighting the importance of oxygen saturation levels and chest CT scans in determining the severity of the disease. Finally, NLR (*n* = 10, 9.8%) was another significant feature, as it is a reliable marker of systemic inflammation and infection (Figure 7a). Regarding performance, the top three ML models were DNN with an average AUC of 0.962, PLS-DA with an average AUC of 0.955, followed by ANN with an average AUC of 0.933 (Figure 7b).

**Figure 7:**
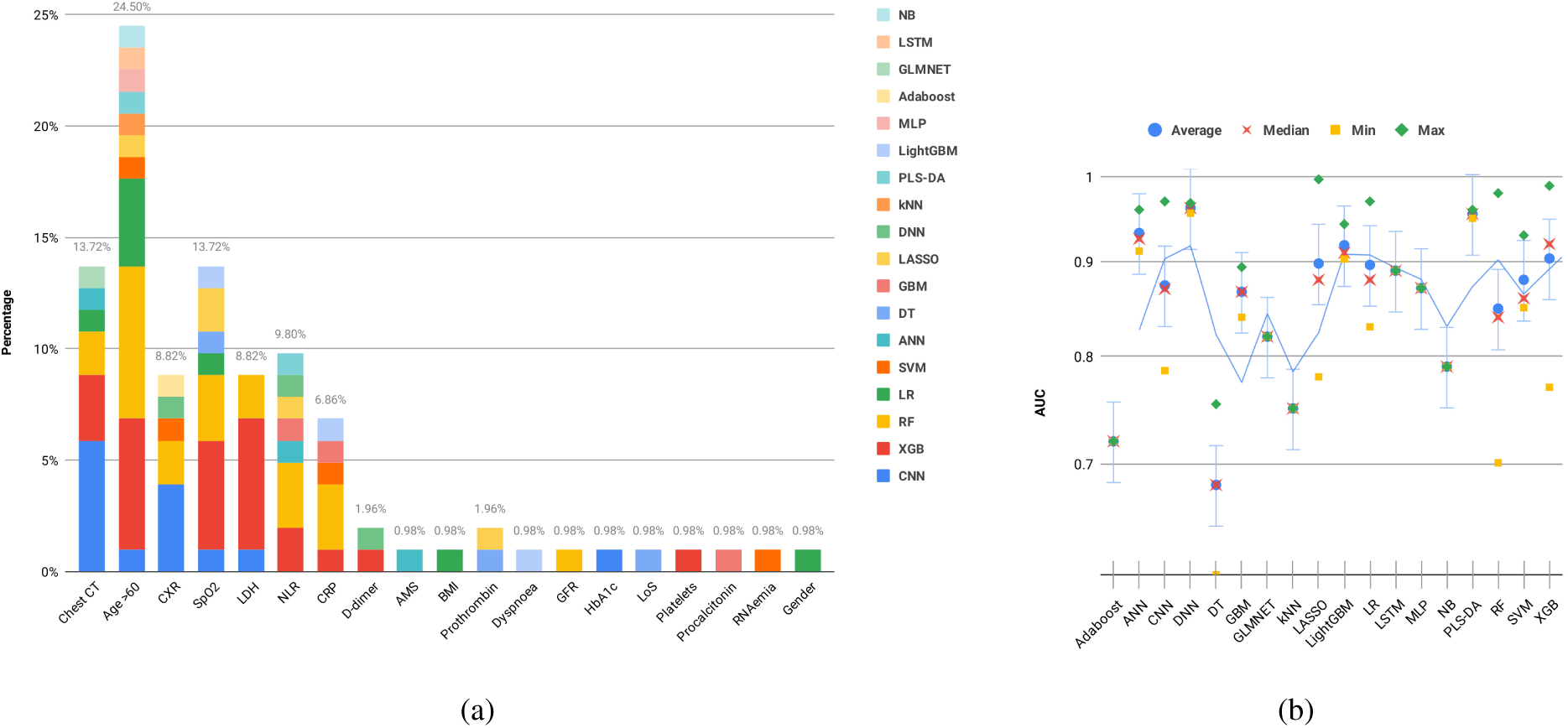
(a) Percentage of machine learning models applied per feature in outcome prediction and (b) Performance metrics for machine learning models in outcome prediction

## 4 Discussion

The COVID-19 pandemic has underscored the need for innovative solutions to alleviate the strain on healthcare systems. Machine learning (ML) holds great promise in the medical field for enhancing the efficiency and accuracy of analysis, facilitating patient risk categorisation, and guiding decision-making. ML algorithms have shown exceptional performance in accurate disease detection, effective severity stratification, and modelling disease prognosis. While numerous studies have explored ML applications for tackling COVID-19 challenges with encouraging results in diagnosis, treatment, prevention, and public health management, many prognostic scores and models suffer from biases due to insufficient methodology, inadequate sample sizes, and limited datasets.

Our study offers a comprehensive systematic literature review of 204 studies investigating the prognostic capabilities of ML models in COVID-19. These studies showcase the versatility of six model categories across five distinct dataset categories, highlighting their potential for broad clinical implementation. The systematic review revealed three primary prognostic themes in the context of COVID-19, with models such as CNN, XGB, and RF featuring prominently. In automated detection and ML-assisted diagnosis, kNN, MLP, and ANN emerged as top-performing models. For severity classification and risk stratification, kNN, ANN, and XGB led the pack. In predicting outcomes and disease progression, DNN, PLS-DA, and ANN were the top performers. These prognostic algorithms are not meant to replace the clinical decision-making process with regard to ascertaining the suitable level of care for patients. Rather, their purpose is to assist healthcare professionals in identifying patients who are at risk of future deterioration.

### 4.1 Prognostic Themes and their Clinical Implications

The clinical implications of using these top-performing models in each theme are multifaceted, with the potential to significantly impact patient care and healthcare system efficiency. Below, we will explore the clinical implications for each theme in more detail.

#### 4.1.1 Automated Detection and AI-assisted Diagnosis

In the realm of automated detection and AI-assisted diagnosis, kNN, MLP, and ANN performed exceptionally well. kNN, or k-Nearest Neighbors, is an intuitive algorithm that classifies new instances by comparing them to their closest neighbors in the feature space. Its success in this theme can be attributed to its simplicity and ability to work well with smaller datasets, which can be common in the early stages of a pandemic. MLP, or Multi-Layer Perceptron, is a feedforward artificial neural network that can model complex relationships between input features and target labels. Its effectiveness may result from its capacity to capture non-linear patterns in the data, which can be crucial for accurate diagnosis. ANN, or Artificial Neural Networks, are inspired by the human brain and excel in pattern recognition and learning from multidimensional data, such as imaging data (CXR and CT scans) commonly used in diagnosis.

The use of kNN, MLP, and ANN in automated detection and diagnosis can lead to earlier identification of COVID-19 cases, particularly in areas with limited access to conventional diagnostic methods such as RT-PCR. These models can assist clinicians in making more informed decisions when evaluating suspected cases, ultimately resulting in quicker isolation and treatment measures. Additionally, the integration of AI into diagnostic processes can reduce the burden on healthcare professionals and minimise diagnostic errors. Moreover, the use of imaging data, such as CXR and CT scans, in conjunction with clinical and laboratory data can help identify high-risk patients, enabling prioritisation of care and resources. Early and accurate diagnosis using these models can also contribute to better public health decision-making and the implementation of effective containment measures.

#### 4.1.2 Severity Classification and Risk Stratification

In the context of severity classification and risk stratification, kNN, ANN, and XGB achieved the best performance. As previously mentioned, kNN’s simplicity and ability to handle small datasets might have contributed to its effectiveness in this theme. ANN’s ability to model complex patterns and relationships also plays a vital role in determining severity and risk. XGB, or eXtreme Gradient Boosting, is an ensemble learning technique that leverages gradient boosting to optimise model performance. Its success may stem from its ability to minimise errors, handle missing values, and automatically manage feature interactions, which are critical when dealing with heterogeneous patient data.

Incorporating kNN, ANN, and XGB models into severity classification and risk stratification can aid healthcare providers in identifying patients at a higher risk of severe illness, complications, or mortality. By accurately classifying patients into predefined risk groups, clinicians can personalise treatment plans and allocate resources more effectively. These models can also help to optimise the use of hospital beds and equipment, such as ventilators, by triaging patients based on their predicted severity. This can lead to better overall patient outcomes, as those requiring more urgent care receive timely attention. Furthermore, identifying high-risk individuals can facilitate the development and evaluation of targeted interventions, such as the use of antiviral medications or immunomodulatory therapies, ultimately improving patient outcomes.

#### 4.1.3 Severity Classification and Risk Stratification

For the prediction of outcomes and disease progression, DNN, PLS-DA, and ANN were the top-performing models. DNN are a type of ANN with multiple hidden layers, which allows them to capture more complex patterns and representations in the data. Their effectiveness in predicting patient outcomes can be attributed to their ability to model hierarchical relationships in data and generalise well to unseen instances. PLS-DA, or Partial Least Squares Discriminant Analysis, is a multivariate technique that combines dimensionality reduction and classification. Its performance may be due to its capability to handle collinear and noisy data, which is common in clinical datasets. ANN, as mentioned earlier, excels in pattern recognition and learning from multidimensional data, which is crucial in predicting disease progression.

Utilising DNN, PLS-DA, and ANN models for predicting patient outcomes and disease progression allows healthcare providers to initiate early interventions and optimise the allocation of healthcare resources. Accurate predictions of patient outcomes, such as ICU admission, mechanical ventilation, and mortality risk, can inform decisions about when and where to deploy interventions, ensuring they are most effective. In addition, these models can support decision-making for patient transfer or referral to specialised care facilities, based on their predicted progression. This can help to balance the load on healthcare systems, enabling more efficient use of resources and better overall patient care.

In summary, the use of top-performing models in each prognostic theme can lead to improvements in diagnostic accuracy, patient risk stratification, and outcome prediction for COVID-19 patients. Integrating these models into clinical practice can optimise healthcare resource allocation, support personalised treatment strategies, and ultimately contribute to better patient outcomes and more efficient healthcare systems.

### 4.2 Feature Selection

The quality of datasets and feature selection strategies are critical for efficient ML and, consequently, precise disease predictions, beyond just the data size. Employing feature selection methods to identify the most informative feature subset for training a model can lead to the development of robust models. Moreover, feature sets comprising histological or pathological evaluations are marked by reproducible values. Given the absence of static entities in the context of clinical variables, it is essential for a ML algorithm to be adaptable to diverse feature sets over time. Feature selection and engineering techniques play a vital role in identifying the most relevant features and transforming them into meaningful variables. This process allows algorithms to decrease the dimensionality of input data, resulting in superior generalisation, improved accuracy, and reduced overfitting [25]. Additionally, feature engineering plays a pivotal role in transforming raw data into informative variables that facilitate ML model training. Expertise within a particular domain is employed to create novel features or adapt existing ones, thereby more effectively encapsulating the inherent patterns present in the data [26]. For example, within the scope of COVID-19 research, scientists have meticulously designed features associated with patients’ clinical, laboratory, and demographic attributes in order to enhance the performance of predictive models [27]. This process of feature engineering ensures the availability of rich, interpretable, and actionable insights for healthcare practitioners. In this systematic review, we analysed the integration of chest imaging, clinical variables, and inflammatory markers to optimise the performance of ML models.

#### 4.2.1 AI-assisted Image Analysis

Recent advances in ML research have highlighted the significance of initial CXR and chest CT findings in the diagnosis of COVID-19. The primary imaging abnormalities identified in COVID-19 positive patients include peripheral ground-glass opacities, bilateral lower lobe consolidation, and diffuse air space disease [28, 29]. In the advanced stages of the illness, less frequent findings consist of bronchiectasis, septal and pleural thickening, and subpleural involvement, which are also evident in other respiratory infections or diseases [28, 29]. Although chest CTs offer greater sensitivity as an imaging technique for the examination of COVID-19 [30], their practicality is constrained by factors such as high radiation doses, limited availability, and substantial operational expenses. CXRs offer rapid visualisation of pathology, their cost-effectiveness, and the portability of imaging equipment is especially beneficial in severe infections. Nonetheless, the typical ground-glass and consolidative changes that are commonly observed in COVID-19 may be challenging to detect on CXRs, limiting their diagnostic value in severe disease [31]. As such, the COVID-19 pandemic has underscored the immense pressure on healthcare systems, including a shortage of trained radiologists. By leveraging AI techniques for automated image analysis, ML models can enhance the speed and accuracy of case identification.

When trained to achieve a level of precision equivalent to that of a radiologist, a ML algorithm has the capacity to deliver results up to approximately 135 times more rapidly and operate continuously, eliminating errors attributable to fatigue [32]. Contemporary research has demonstrated the potential of AI-driven applications in identifying COVID-19 through chest CT scans and distinguishing it from other conditions, such as pneumonia [33, 32]. These applications employ deep learning models to analyse medical images effectively, achieving high accuracy and AUC scores. Verma et al. developed an AI-driven Android application achieved an impressive AUC of 0.999 and accuracy of 97.93% for detecting COVID-19 from chest CT scans [33]. Similarly, Li et al. developed an AI system to differentiate COVID-19 from community-acquired pneumonia using chest CT scans. The AI system demonstrated a high accuracy of 90.6% and an AUC of 0.96 in the first test set. In the second test set, the AI system achieved an accuracy of 87.5% and an AUC of 0.95 [32].

Moreover, deep learning-based techniques using CNNs have shown promise in the identification and classification of pneumonia from CXR images, achieving high accuracy rates of 92.1% and AUC values of 0.967, as observed in the works of Elshennawy and Ibrahim [34]. In addition, Zhong and colleagues have showcased the capability of CNNs to not only retrieve, but also rank comparable CXRs by evaluating their radiographic similarities with an AUC score of 0.913 [35]. Guefrechi et al. introduced a deep learning-based approach to diagnose COVID-19 from CXR images. The top-performing model, a modified DenseNet-169 architecture, attained an overall accuracy of 97.3%, sensitivity of 98.3%, specificity of 96.3%, and an AUC of 0.993 for detecting COVID-19 cases from CXR images [36].

Balaha et al. proposed a comprehensive framework for the accurate recognition and prognosis of COVID-19 patients using a deep transfer learning and feature classification approach. By combining CNNs with ML classifiers, their framework analysed medical images and extracted crucial features for classification, resulting in an overall accuracy of 96.9%, sensitivity of 98.1%, specificity of 95.6%, and an AUC of 0.969 for the prognosis task [37]. In a related study, Manokaran et al. employed transfer learning with four pre-trained CNNs for COVID-19 detection through CXRs. Their best-performing model, Inception-ResNetV2, achieved 98.2% accuracy, 96.3% sensitivity, 99.4% specificity, and an AUC of 0.995 [38].

Accurate identification of these subtle radiological manifestations and patterns enhances COVID-19 detection by quantifying lung involvement and abnormalities in CT and CXR images. These systems showcase the potential for clinical implementation in managing COVID-19 patients, providing healthcare professionals with a rapid diagnostic tool and means to assess of disease progression.

#### 4.2.2 Bedside Prognosticators: Age, SpO2, and Cough Signals

As the COVID-19 pandemic continues to challenge healthcare systems worldwide, understanding the role of key clinical predictors, such as age, oxygen saturation levels, and cough signals, is crucial in detecting and managing the disease and its progression.

Advanced age has been recognised as a significant risk factor and an important predictive feature associated with COVID-19 severity and disease progression. Elderly patients, typically defined as those 60 years and older, have been observed to experience a higher incidence of respiratory failure and necessitate more prolonged treatment regimens in contrast to their younger counterparts [39], implying that the geriatric population may exhibit a less favourable response to therapeutic interventions. Older individuals are also more prone to co-morbidities, which are recognised as independent risk factors for COVID-19 prognosis. In order to better evaluate a patient’s risk profile, ML algorithms have been developed that can utilise age as either a categorical or continuous variable. By incorporating age in these models, healthcare providers can more accurately identify patients who may require more intensive interventions or monitoring. This, in turn, could lead to improved outcomes for older individuals afflicted with COVID-19 [40].

Additionally, prior studies have revealed a direct relationship between advanced age and elevated mortality rates among hospitalised patients with COVID-19 [41]. The increased vulnerability to infections observed among the elderly population can be attributed to the diminished expression of type I interferon-beta, leading to an atypical reaction to viral pathogens [42]. Furthermore, age-related lymphocyte function impairment, coupled with abnormal type 2 cytokine expression results in prolonged pro-inflammatory responses. This compromised host response to viral replication leads to suboptimal clinical outcomes and increased mortality rates [42].

Roimi et al. developed and validated a ML model to forecast the illness trajectory and hospital utilisation of COVID-19 patients using nationwide data [43]. By incorporating age as a pivotal predictive factor, the model could more accurately identify patients at higher risk of severe illness and prioritise their care. This would, in turn, enable healthcare systems to allocate resources more effectively and improve patient outcomes. In a parallel study conducted by An et al., researchers devised an ML model to prognosticate COVID-19 mortality, drawing upon data from a nationwide Korean cohort. The gradient boosting machine model demonstrated superior performance, achieving an AUC of 0.938, which signifies exceptional accuracy in predicting mortality. Age emerged as the foremost predictive factor for COVID-19 mortality, placing older patients at heightened risk [44].

In contrast to conventional pneumonia, the initial phases of COVID-19 exhibit minimal discernible symptoms, such as dyspnoea. This occurs as carbon dioxide exchange via alveoli remains unimpeded during these stages. Nevertheless, alveolar collapse disrupts oxygen exchange, culminating in “silent hypoxia,” which causes pneumonia progression in the absence of clinical manifestations. This hypoxic state stimulates local inflammatory markers, leading to further damage and exacerbating hypoxia [45]. Consequently, low oxygen levels may serve as a critical determinant in revealing pneumonia progression and assessing the severity of patients’ conditions.

Vital signs, particularly oxygen saturation levels, serve as critical indicators for severe disease. SpO2 is a non-invasive and effective method for evaluating a patient’s respiratory function, offering valuable insights into patients’ oxygenation status and hypoxia. Therefore, low levels of oxygen saturations reflect underlying disease severity, the necessity for cardiorespiratory resuscitation, and the potential risk of future decompensation in the absence of adequate medical intervention. ML models have the potential to integrate non-invasive parameters such as age and SpO2 data alongside other clinical parameters to categorise patients according to illness severity, forecast disease progression, and monitor treatment response [46]. Several studies, such as the one conducted by Calvillo-Batllés et al., have developed prediction models to evaluate the severity and mortality risk of COVID-19 patients by analysing a dataset encompassing clinical, laboratory, and imaging data from COVID-19 patients. A key predictive factor identified in this study was SpO2, which demonstrated an impressive AUC of 0.97 [47]. The research highlights the benefits of utilising a combination of data sources, including SpO2, to improve the accuracy of prediction models for COVID-19 patients, ultimately enabling clinicians to make well-informed treatment decisions in emergency departments. In a separate study by Kocadagli, the authors developed a hybrid model combining decision tree-based and deep learning models for prognosis evaluation. Applied to a dataset of 1,125 COVID-19 patients, the model identified important risk factors, with SpO2 being highly predictive of poor prognosis and achieved a high AUC of 0.98 [48].

This information is essential for guiding appropriate interventions, for instance, a patient experiencing a rapid decline in SpO2 levels might necessitate prompt immediate hospitalisation, oxygen therapy, or mechanical ventilation to avert additional complications [45]. Furthermore, continuous monitoring of SpO2 levels can be instrumental in assessing the efficacy of various treatments, such as antiviral medications, corticosteroids, or supplemental oxygen. By monitoring SpO2 fluctuations over time, medical professionals can evaluate the outcomes of the implemented therapeutic interventions and modify their strategies accordingly [41]. Apart from its application in managing individual patients, SpO2 data can also be harnessed for large-scale epidemiological studies to better understand the clinical course of COVID-19. Through the examination of SpO2 values across diverse populations, investigators can discern trends and patterns, thereby facilitating the implementation of more effective therapeutic interventions and public health policies [39].

Recent literature has highlighted the significant potential of innovative approaches for detecting COVID-19 through the analysis of cough audio signals. This diagnostic method employs ML classifiers in conjunction with acoustic biomarker feature extraction techniques to preliminarily screen for COVID-19 from cough recordings, offering a non-invasive, real-time pre-screening technique [49]. As one of the most prevalent and notorious symptoms of COVID-19, cough audio analysis can supplement existing disease containment measures in regions with low infection rates and mitigate the impact in areas experiencing high infection rates, where asymptomatic individuals may unknowingly transmit the virus.

Researchers such as Hossain and colleagues have proposed signal processing and ML techniques to analyse cough signals and identify unique features related to COVID-19 infection [50]. Their automated approach, called *Covidenvelope*, demonstrated a classification accuracy of 95.10%, a sensitivity of 94.64%, and a specificity of 96.32% for COVID-19 diagnosis. Another notable study by Laguarta et al. devised an artificial intelligence framework that generates personalised patient saliency maps, enabling longitudinal patient monitoring in real-time, non-invasively, and at virtually no expense [51]. Upon validation with patients diagnosed through formal testing methods, this algorithm exhibits a COVID-19 sensitivity of 98.5%, a specificity of 94.2%, and an AUC of 0.97.

These findings suggest that AI-driven cough recording analysis can function as a rapid, non-invasive, and cost-effective solution for large-scale COVID-19 asymptomatic screening, augmenting existing efforts to control virus transmission. Nevertheless, it is essential to conduct further validation using larger and more diverse datasets to enhance generalisability prior to implementing this approach in real-world settings. Ultimately, by incorporating age, SpO2, and cough signals in ML algorithms, healthcare professionals can enhance patient care, inform treatment decisions, and optimise resources, ultimately contributing to a more efficient and targeted response to the pandemic.

#### 4.2.3 Serum Inflammatory Biomarkers

Inflammatory markers, such as lactate dehydrogenase (LDH), neutrophil-to-lymphocyte ratio (NLR), and C-reactive protein (CRP), are valuable indicators of a patient’s inflammatory and overall physiological state. ML models can leverage these biomarkers, along with other clinical features, to stratify patients by severity, predict outcomes, and identify potential complications [52].

LDH, an intracellular enzyme predominantly found in lung tissue (isozyme 3), is released into circulation during severe infections as a result of cytokine-mediated tissue damage [53]. This release of LDH in high-risk COVID-19 patients suggests its potential utility as a marker for severe disease. Elevated LDH levels may indicate lung tissue injury and inflammation caused by SARS-CoV-2 infection and concurrent lung fibrosis. Furthermore, a robust immune response and the ensuing cytokine storm, a massive, dysregulated release of pro-inflammatory cytokines and chemokines, may lead to multi-organ damage and subsequent elevations in LDH levels [54]. Severe SARS-CoV-2 infection-induced inflammation can cause endothelial damage, disruption of coagulation cascade function, and coagulopathy. Additionally, high LDH levels have been associated with co-infections and could serve as potential prognostic indicators in severe bacterial infections. Numerous studies have investigated the role of LDH in COVID-19 disease progression, linking elevated LDH levels to increased risk, severity, and fatality [53, 54, 40, 55].

Neutrophils and lymphocytes, subsets of white blood cells, play critical roles in the inflammatory response and immune system, respectively. The Neutrophil-to-Lymphocyte Ratio (NLR), a readily calculated inflammatory marker, has been recognised as an independent risk factor for severe illness in COVID-19 patients, with low lymphocyte and high neutrophil counts being predictive of ICU admission. Several studies have corroborated these findings, highlighting NLR as a practical and important prognostic factor derived from routine blood tests [56, 57, 55]. COVID-19 severity is primarily influenced by the body’s innate inflammatory response, with more severe cases often attributed to cytokine storms [58]. This reaction, associated with severe COVID-19 cases, may result in multi-organ failure and death, emphasising the complex interaction between the host immune system and the virus. Assessing neutrophil and lymphocyte levels upon hospital admission can facilitate early risk stratification, enabling healthcare professionals to identify and prioritise patients exhibiting higher inflammation levels as severe COVID-19 cases.

Serum biomarkers, such as inflammatory and acute phase reactants, have been extensively studied for their potential to offer early clinical indications of severe disease onset in COVID-19 patients [59, 60]. CRP, a widely used marker of inflammation, has been identified as a robust prognosticator of disease severity and poorer prognosis in COVID-19 cases [59]. Elevated CRP levels have also been significantly correlated with the extent of pulmonary involvement and the size of lung lesions in CT imaging [60], suggesting CRP as a valuable biomarker for disease severity and potential prognostic feature for respiratory failure during early stages of COVID-19. Elevated levels of high-sensitivity C-reactive protein (hs-CRP) have been observed to exhibit an inverse relationship with pulmonary function. Extremely high CRP levels may signify the development of severe bacterial superinfections, a common complication in critically ill COVID-19 patients and a potential contributing factor to heightened mortality rates [61]. Early identification of patients at higher risk for superinfections or other complications, before significant increases in CRP and LDH levels, could facilitate more efficient treatment strategies and improve patient outcomes.

ML models incorporating these inflammatory markers have demonstrated robust performance in predicting COVID-19 mortality. For instance, Booth et al. developed a prognostic model that predicted mortality up to 48 hours before death with an AUC of 0.93, surpassing traditional logistic regression models [62]. Zhu et al. created a deep-learning model with an accuracy of 93.1%, sensitivity of 89.3%, specificity of 95.6%, and an AUC of 0.97. They identified D-dimer, O2 Index, NLR, CRP, and LDH as the top 5 predictors of COVID-19 mortality [63]. Furthermore, Yan et al. developed an interpretable mortality prediction model with an AUC of 0.94, accurately predicting mortality more than ten days in advance with an accuracy exceeding 90% [40]. Lastly, Karthikeyan et al. established a ML-driven clinical decision support system aimed at predicting COVID-19 mortality in the initial stages, achieving an AUC of 0.979, sensitivity of 95.3%, specificity of 92.9%, and accuracy of 94.1% using NLR, LDH, CRP, and age as predictive biomarkers [55]. These findings underscore the prognostic potential of ML models that incorporate inflammatory markers in assisting clinicians with informed treatment decisions, ultimately improving patient outcomes in COVID-19 cases.

The research by Schöning et al., Alle et al., and Altini et al. underscores the effectiveness of ML models in stratifying COVID-19 severity using various clinical data [64, 65, 66]. Schöning et al. developed the COSA score and compared it to ML models, with the gradient boosting machine model exhibiting the highest performance and an AUC of 0.960 [64]. Alle et al. utilised an ensemble model consisting of gradient boosting machines, random forests, and LASSO logistic regression, achieving an AUC of 0.91, sensitivity of 0.81, and specificity of 0.88 [65]. Finally, Altini et al. developed models based on hematochemical parameters, with the extreme gradient boosting model yielding the highest accuracy [66]. These models, particularly ensemble models, have demonstrated high accuracy and AUC values in predicting patient outcomes, such as recovery, ICU admission, and death. This research highlights the potential of ensemble models in risk classification, contributing to improved patient outcomes and more efficient management of the ongoing pandemic.

### 4.3 Machine Learning Techniques

A critical aspect of optimising ML models for COVID-19 prognostication lies in selecting the most appropriate algorithms. A wide range of algorithms, including neural networks and ensemble models have been employed in the literature to tackle various COVID-19-related tasks [67, 68]. The choice of the most suitable algorithm is contingent upon variables including the dimensionality and complexity of the dataset, as well as the specific problem being addressed. Undertaking a comparative analysis of these algorithms is vital to pinpoint their respective strengths and weaknesses in the context of COVID-19 prognostication, ensuring the adoption of the most effective solution [68].

#### 4.3.1 Neural Networks

In the realm of COVID-19 prognostication, neural networks have emerged as a powerful tool for predicting disease outcomes and progression. Among various ML algorithms, neural networks, particularly deep learning models, have demonstrated their ability to effectively process complex and high-dimensional data, such as medical images and electronic health records [69, 70]. These models can capture intricate relationships and nonlinear patterns in the data, which contributes to their robust performance in a variety of COVID-19-related tasks [67].

One successful application of neural networks in COVID-19 prognostication has been Convolutional Neural Networks due to their ability to process and analyse image data. Their hierarchical structure and specialised layers make them well-suited for medical imaging applications, including the diagnosis and analysis of COVID-19 using chest X-ray and CT scan images [71, 32, 72]. CNNs automatically learn relevant features from raw image data, eliminating the need for manual feature engineering allowing researchers to develop models for detecting COVID-19 in medical images [73]. For example, Wang et al. used a CNN to distinguish COVID-19 pneumonia from other types of pneumonia with chest X-ray images, achieving 94% accuracy [74].

In addition to chest X-ray images, CNNs have been applied to analyse chest CT scans for COVID-19 detection and assessment. Li et al. employed a deep learning model called COVNet to differentiate COVID-19 from other types of pneumonia in chest CT scans, achieving an AUC of 0.96 [32]. Moreover, CNNs have been used to assess disease severity and predict patient outcomes. In a study by Shi et al., a CNN-based model quantified lung abnormalities in COVID-19 patients using chest CT scans, demonstrating a high correlation with radiologist assessments and showing promising results in detecting the disease and analysing disease progression [75].

Artificial Neural Networks constitute another important algorithm, demonstrating considerable potential in COVID-19 prognostication due to their capacity to recognise and model intricate patterns within datasets. Numerous studies have illustrated the efficacy of ANNs in predicting COVID-19 presence using clinical and laboratory data, as well as forecasting the risk of COVID-19-associated mortality based on demographic and clinical information [76, 77]. Moreover, these networks have been utilised to model disease progression and assess the effectiveness of interventions. For example, El Amine et al. employed an ANN to predict the severity of COVID-19 infection using chest CT images, achieving an accuracy of 88.8% [78]. Furthermore, Pourhomayoun et al. implemented an ANN to estimate COVID-19 hospitalisation, ICU admission, and mortality, attaining an AUC of 0.83, 0.86, and 0.79, respectively [79].

Deep Neural Networks have shown potential in COVID-19 prognostication due to their capacity to process large and complex datasets [69]. As a subtype of ANNs, DNNs employ multiple layers of neurons to learn hierarchical representations of input data, thereby facilitating the learning of increasingly abstract and complex features. These models have been employed in tasks such as diagnosis, disease detection, severity stratification, and prognosis modelling. For example, Xu et al. utilised a DNN to predict COVID-19 severity based on chest CT images with 85.7% accuracy [80], while Alqudah et al. achieved 95.6% accuracy in detecting COVID-19 from chest X-ray images [81]. In a similar vein, in a study conducted by Ahmad et al., a DNN-based model was designed to estimate the risk of COVID-19 mortality using demographic and clinical data, reaching an accuracy of 94.1% [82]. Likewise, Miotto et al. employed a DNN to forecast COVID-19 patient outcomes, demonstrating enhanced predictive performance in comparison to conventional regression models [70].

Neural networks, including deep learning models such as CNNs, ANNs, and DNNs, have each exhibited their unique strengths in handling complex and high-dimensional data. They have proven to be invaluable tools in the field of COVID-19 prognostication, demonstrating remarkable success in predicting disease outcomes, progression, and response to interventions. By capitalising on the capacity of these models to discern intricate relationships and nonlinear patterns, healthcare providers can make more informed, data-driven decisions when managing patients, ultimately leading to better patient outcomes and more efficient allocation of resources during the ongoing pandemic.

#### 4.3.2 Ensemble Models

Ensemble models have been gaining traction in the field of medical research, particularly in the context of COVID-19, due to their ability to harness the power of multiple ML algorithms for improved predictive performance. Ensemble models, which combine different ML algorithms like CNNs, XGB, and ANNs, have the potential to significantly enhance prognostic model performance [83]. These models work by aggregating the outputs of multiple base models, effectively capitalising on their individual strengths while compensating for their weaknesses. In doing so, ensemble models can yield more accurate and reliable predictions, which are critical for effective disease detection, severity stratification, and prognosis modelling for COVID-19 [84, 64]. In turn, this enables healthcare providers to make more informed, data-driven decisions when managing COVID-19 patients, leading to better patient outcomes and more efficient allocation of resources.

Various ensemble techniques have been employed in COVID-19 research, each with their unique advantages and challenges. One popular ensemble technique is bagging (bootstrap aggregating), which involves training multiple base models on random subsets of the training data and aggregating their predictions through a majority vote or averaging [85]. Bagging can effectively reduce overfitting and improve stability in the predictions, as demonstrated by a study conducted by Sharma et al., which used bagging with decision trees to predict COVID-19 infection and achieved an accuracy of 93.7% [86]. This approach was also successfully applied by Al-Waisy et al., who developed a bagged random forest model to predict COVID-19 mortality, achieving an AUC of 0.91 [87].

Boosting is another ensemble technique where base models are trained sequentially, with each new model aiming to correct the errors of its predecessor [88]. This approach has been employed in the development of the well-known XGB algorithm [89]. In a study by Altini et al., an XGB model was used to predict COVID-19 prognosis based on haematochemical parameters, achieving the highest accuracy with an AUC of 0.85 for recovery, 0.87 for ICU admission, and 0.89 for death [90]. Similarly, Nascimento et al. applied an XGB model to predict COVID-19 severity using clinical and demographic data, resulting in an AUC of 0.88 [91].

Stacking, or stacked generalisation, is a third ensemble technique that involves training a higher-level model, known as a meta-model, on the outputs of multiple base models [92]. This approach allows for more complex and sophisticated decision-making processes, as the meta-model can learn to optimally combine the predictions of the base models [93]. A study by Zhang et al. employed a stacking ensemble of deep learning models, including CNNs and recurrent neural networks (RNNs), for COVID-19 detection using CXR images [94]. Their model achieved an AUC of 0.98, outperforming the base models. Additionally, Ozturk et al. developed a stacked ensemble model combining CNNs and XGB for COVID-19 detection from chest CT scans, reaching an AUC of 0.996 [95].

Ensemble models can also be tailored to specific clinical settings and patient populations, as shown by a study conducted by Gupta et al. [96]. The researchers developed an ensemble model using CNNs, LSTM networks, and decision trees to predict COVID-19 severity in paediatric patients. The model demonstrated high predictive performance with an AUC of 0.92, enabling early identification and intervention for high-risk children. In another study, Punn et al. designed an ensemble model combining CNNs and LSTMs to predict COVID-19 severity in elderly patients [97]. This model achieved an AUC of 0.94, demonstrating its effectiveness in identifying high-risk older adults who may require targeted interventions.

Furthermore, ensemble models can be used to incorporate diverse data types and sources, such as demographic information, clinical measurements, imaging data, and omics data, into a single predictive model [98]. In a study by Pereira et al., an ensemble model that combined clinical, demographic, and radiomic features was developed to predict COVID-19 severity [99]. The model achieved an AUC of 0.89, highlighting the benefits of integrating multiple data sources for improved prognostication. Similarly, Duarte et al. created an ensemble model incorporating demographic data, clinical measurements, and viral load information to predict the need for ICU admission in COVID-19 patients, obtaining an AUC of 0.93 [100].

Moreover, ensemble models can effectively capture the intricate relationships and nonlinearities within the data, allowing for a more comprehensive representation of the underlying structure. Additionally, they can provide a higher degree of model robustness, as they are less susceptible to overfitting and can adapt better to new data [83]. The utilisation of ensemble models in the context of COVID-19 prognostication is a promising avenue for advancing disease management strategies, offering enhanced predictive performance through the combination of multiple base models. These models have been successfully applied to a wide range of tasks, including disease detection, severity stratification, and prognosis modelling [86, 90, 94]. Moreover, ensemble models can be tailored to specific clinical settings and patient populations, as well as incorporate diverse data types and sources for more comprehensive predictions [96, 99]. As the COVID-19 pandemic continues to evolve, ensemble models are poised to play a crucial role in improving our understanding of the disease and informing clinical decision-making.

### 4.4 Challenges and Limitations

Several challenges and limitations affect the implementation and generalisability of ML models in COVID-19 prognosis, including biases due to insufficient reporting, methodology, inadequate sample sizes, and limited datasets. The lack of standardisation across studies further complicates the comparison and validation of different ML models.

#### 4.4.1 Sources of Bias

Despite promising results, multiple obstacles and limitations influence the implementation and generalisability of ML models in COVID-19 prognosis. Challenges include biases originating from inadequate reporting, methodological issues, insufficient sample sizes, and limited datasets. The absence of standardisation among studies complicates the difficulty in comparing and validating distinct ML models. Another significant constraint is overfitting, where ML models may excel on the training dataset but struggle to generalise to novel, unobserved data. This problem can result in overly optimistic performance metrics and hinder the practical applicability of ML models in real-world settings.

Moreover, only a few authors have mentioned the actual integration of ML into clinical workflows. This systematic review presents a reasonable representation of the latest trends in clinical applications of ML; however, it may not encompass the complete potential or actual applications. As a result, selection bias is unavoidable, and publication bias cannot be excluded. While AI is lauded for its contributions to cost reduction, enhanced efficiency, throughput, accuracy, and precision, issues surrounding reliability, ethics, transparency, and generalisability have emerged. Growing ethical debates and concerns may impede mainstream adoption in clinical practice. Algorithmic bias is an omnipresent risk, highlighting the need for high-quality data collection.

#### 4.4.2 Standardisation

To ensure the reliability and applicability of ML models in COVID-19 prognosis, several factors must be taken into consideration. In particular, standardisation, validation, and reproducibility in ML studies are crucial. Standardisation is critical when it comes to data collection, pre-processing, and reporting. By doing this, it enhances algorithm comparability and facilitates the identification of ideal approaches. It is also worth noting that external validation using independent datasets is equally critical for assessing the generalisability and performance of ML models in diverse healthcare settings.

Reproducibility is another essential component of scientific research, particularly in ML studies. Even minor adjustments in parameters or training data can lead to substantial performance variations. To ensure reproducibility in ML studies, it remains crucial to report methodologies, data pre-processing techniques, and model architectures transparently. To elaborate further, it may be helpful to provide detailed descriptions of the dataset, including the sample size, data source, and data types. Similarly, it may be necessary to explain the feature selection process and the rationale behind it. By doing so, it increases the transparency and reproducibility of the study, which is critical when it comes to scientific research.

#### 4.4.3 Ethical Considerations and Data Privacy

As the adoption of ML models in healthcare continues to expand, addressing ethical considerations and data privacy concerns becomes crucial in maintaining a scientific and professional approach. The utilisation of patient data in AI studies raises questions regarding data security, informed consent and the potential biases embedded within algorithms. To maintain public trust and promote widespread adoption of these advanced technologies, it is imperative to protect patient privacy and ensure responsible development and deployment of such models.

A viable approach to foster responsible development and deployment is the establishment of comprehensive ethical guidelines. These guidelines should outline the proper use of patient data and emphasise the need for transparency during algorithm development. Collaboration between healthcare providers and data scientists is essential in identifying potential biases in algorithms and creating protocols to address any emerging issues. This cooperative effort can contribute to a more equitable and unbiased implementation of ML in healthcare.

By addressing ethical considerations and data privacy concerns collectively, we can work towards ensuring the safe, effective and extensive adoption of ML in healthcare. Such an approach not only benefits the scientific community but also has the potential to improve patient outcomes and overall healthcare efficiency.

### 4.5 Future Directions

As the use of ML models in COVID-19 prognostication and management continues to evolve, several key areas warrant further investigation to ensure their successful implementation and widespread adoption.

#### 4.5.1 Model Optimisation

The process of optimising hyperparameters and feature selection is a crucial aspect of ML model development for healthcare applications, as it directly impacts model performance, complexity, and the risk of overfitting [101]. Overfitting occurs when a model becomes too tailored to the training data, reducing its ability to generalise to new, unobserved data. To counter overfitting, selecting suitable ML models for distinct clinical situations is important, with hyperparameter tuning and feature selection serving as key factors in achieving a balance between performance and complexity. Careful consideration of generalisability, interpretability, and ease of deployment during model selection ensures that the chosen models are best suited for their intended clinical scenarios [102].

#### 4.5.2 Integration into Clinical Decision-making

Ongoing research and development in ML for COVID-19 prognostication and management is of paramount importance. This is particularly true in light of the emergence of new variants, the ever-changing healthcare demands, and the continuous integration of novel insights pertaining to the disease. Identifying opportunities to integrate ML models into existing clinical decision-making processes, such as electronic health record systems, clinical decision support tools, and telemedicine platforms, is of utmost importance. Strategies to facilitate implementation and adoption by healthcare providers should address barriers related to technical expertise, stakeholder engagement, and ethical considerations.

#### 4.5.3 Interdisciplinary Collaboration and Open Science

Although AI serves as a powerful tool in supporting diagnosis and prognosis, its effectiveness relies on factors such as appropriate infrastructure, unbiased data sharing, and translation of models into usable solutions. To discover comprehensive solutions for combating COVID-19, international cross-disciplinary collaborations are imperative for carefully identifying time, course, and region-specific clinical actions in response to the pandemic. Future research endeavours should leverage data from national and international collaborative COVID-19 repositories, providing larger and more diverse datasets. Additionally, implementing decentralised AI architectures can address concerns regarding the sharing of sensitive clinical data. These systems allow the creation and training of robust ML models without directly exchanging patient information, thereby safeguarding patient privacy.

#### 4.5.4 Addressing Research Gaps

Further investigation should target research gaps and areas of potential advancement, including the development of ML models for predicting long-term outcomes, post-acute sequelae of COVID-19 and the effectiveness of therapeutic interventions. Additionally, addressing ethical concerns, transparency and generalisability of ML models is essential to promote their mainstream adoption in clinical practice and foster a more cohesive and effective approach to harnessing the potential of AI in managing and treating COVID-19.

## 5 Conclusion

The COVID-19 pandemic has underscored the potential of machine learning (ML) models to transform healthcare systems and guide data-driven decision-making. Our comprehensive systematic literature review of 204 studies revealed three primary prognostic themes in the context of COVID-19: automated detection and AI-assisted diagnosis, severity classification and risk stratification, and prediction of outcome and disease progression. ML models have shown exceptional promise in analysing chest imaging, utilising age and oxygen saturation as critical predictive features, and incorporating serum inflammatory biomarkers to predict disease outcomes. Ensemble models, in particular, have emerged as a powerful tool in medical research capable of integrating multiple ML algorithms for improved predictive performance, ultimately leading to better patient outcomes and more efficient allocation of resources.

However, further research, validation, and standardisation are necessary to establish reliable AI-based diagnostic tools and improve disease detection and management. As the global scientific community grapples with COVID-19, optimising ML models for diagnostic purposes, disease classification, and prognostic predictions is vital in enhancing our comprehension of the disease and informing clinical decision-making. Advancements in feature selection, engineering, algorithm choice, ethical considerations, model evaluation, and integration with other technologies will lead to more effective solutions. By leveraging the wealth of available data and harnessing the power of ML, researchers and healthcare professionals can better anticipate disease outcomes, optimise patient management, and improve the overall response to the COVID-19 pandemic. These findings can enhance the efficiency of healthcare workers in triaging patients, improve patient outcomes, and assist policymakers in allocating limited healthcare resources and integrating public health efforts aimed at combating the devastating effects of the pandemic and beyond.

## Data Availability

All data produced in the present study are available upon reasonable request to the authors

## Notes

### Competing Interest Statement

The authors have declared no competing interest.

### Funding Statement

This study did not receive any funding

### Author Declarations

It is a systematic review paper.

